# Importation of SARS-CoV-2 following the *“semaine de relâche”* and Québec’s COVID-19 burden - a mathematical modeling study

**DOI:** 10.1101/2020.07.20.20158451

**Authors:** A Godin, Y Xia, DL Buckeridge, S Mishra, D Douwes-Schultz, Y Shen, M Lavigne, M Drolet, AM Schmidt, M Brisson, M Maheu-Giroux

## Abstract

**Background:** The Canadian epidemics of COVID-19 exhibit distinct early trajectories, with Québec bearing a very high initial burden. The *semaine de relâche*, or March break, took place two weeks earlier in Québec as compared to the rest of Canada. This event may have played a role in the spread of severe acute respiratory syndrome coronavirus 2 (SARS-CoV-2). We aimed to examine the role of case importation in the early transmission dynamics of SARS-CoV-2 in Québec.

**Methods:** Using detailed surveillance data, we developed and calibrated a deterministic SEIR-type compartmental model of SARS-CoV-2 transmission. We explored the impact of altering the number of imported cases on hospitalizations. Specifically, we investigated scenarios without case importation after March break, and as scenarios where cases were imported with the same frequency/timing as neighboring Ontario.

**Results:** A total of 1,544 and 1,150 returning travelers were laboratory-confirmed in Québec and Ontario, respectively (with symptoms onset before 2020-03-25). The cumulative number of hospitalizations could have been reduced by 55% (95%CrI: 51-59%) had no cases been imported after Québec’s March break. However, had Québec experienced Ontario’s number of imported cases, cumulative hospitalizations would have only been reduced by 12% (95%CrI: 8-16%).

**Interpretation:** Our results suggest that case importation played an important role in the early spread of COVID-19 in Québec. Yet, heavy importation of SARS-CoV-2 in early March could be insufficient to resolve interprovincial heterogeneities in cumulative hospitalisations. The importance of other factors-public health preparedness, responses, and capacity-should be investigated.

## Introduction

The Canadian epidemic of coronavirus disease 2019 (COVID-19) is marked by stark geographic heterogeneities (1). Despite reporting its first case on February 28, 2020 — close to a month after Ontario on January 25^th^ and British Columbia on January 28^th^ — Québec quickly became the epicenter of the Canadian COVID-19 epidemic. The disease’s mortality burden in that province, at 653 per million population, is 3.5 times as high as neighboring Ontario (186 per million) and 19 times that of British Columbia (at 35 per million) (2). The death toll in Québec is on par with that of some of the worst affected European countries. The province also has one of the highest ratios of confirmed cases per million population in the world (6,540 per million), higher than France (2,560 per million), Spain (6,370 per million), the United Kingdom (4,200 per million), and only recently surpassed by the United States (Figure 1)(3) - albeit these comparisons are caveated by underlying testing efforts.

**Figure 1.**
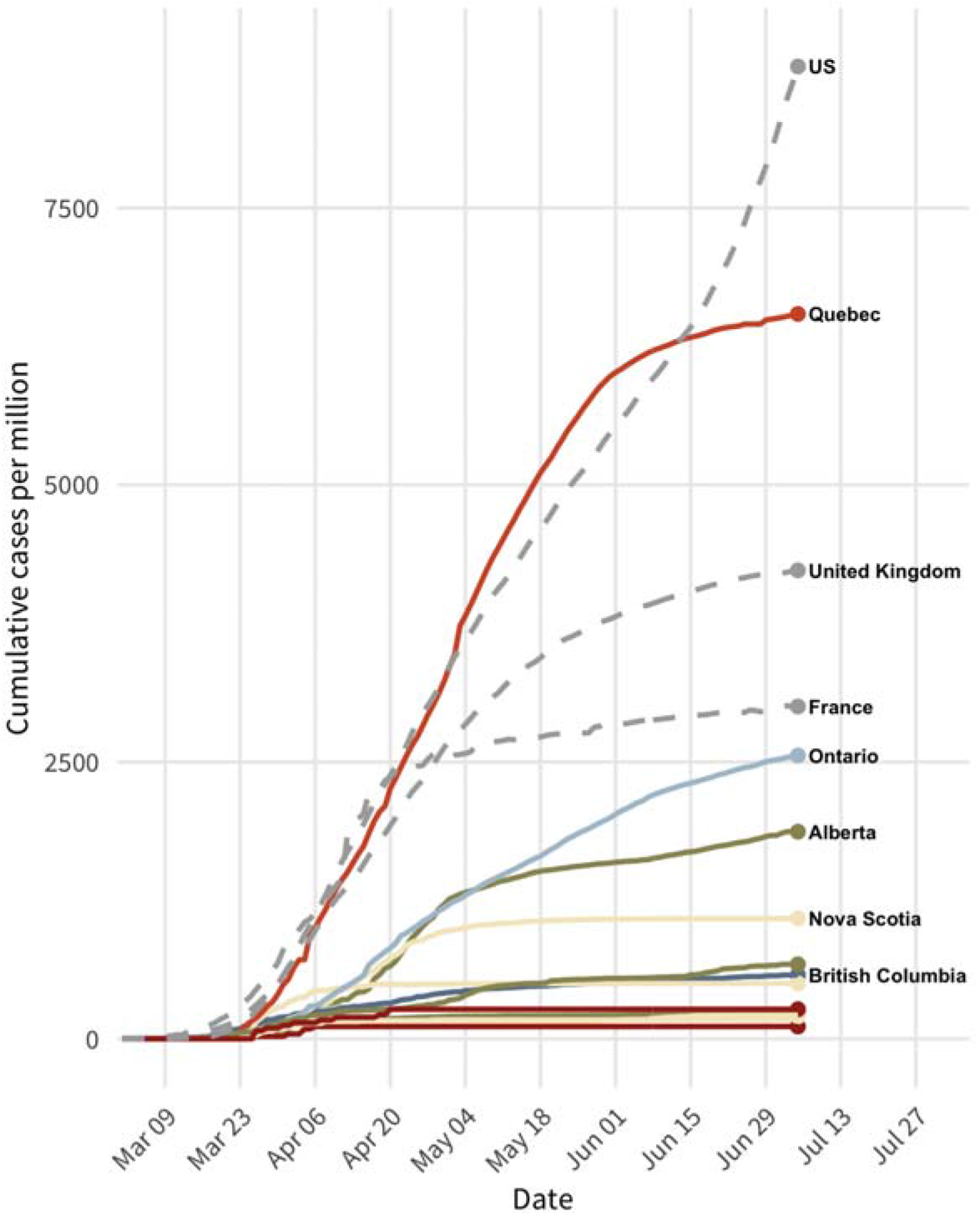
Cumulative confirmed (diagnosed) cases per million population in Canadian provinces and territories and selected countries as of July 6^th^ 2020 (3).

Within Canada, such interprovincial epidemic differences are puzzling given general similarities in age structure, health systems, and timing of local physical distancing measures. For example, British Columbia and Ontario’s ratios of confirmed cases per population are 11 and 2.6 times lower, respectively, than that recorded in Québec (as of July 5^th^ 2020). One hypothesis for these differences, is that the *semaine de relâche*, or March break, played a role in seeding the Québec epidemic. This March break took place during the week of March 2^nd^ and, for some schools, the week of March 9^th^. This event, up to 2-weeks earlier in Québec compared to other Canadian provinces, could have led to a sudden and high number of imported cases that contributed to onward transmission —quickly overwhelming Québec’s capacity for testing and contact tracing— at a time when physical distancing measures had yet to be enacted and implemented. In other provinces, public health authorities recommended against travel during their March break (4,5).

A systematic examination of the impact of imported cases on COVID-19 burden in Québec has yet to be conducted. In this study, we analyze and compare surveillance data from Québec and Ontario regarding the number and timing of imported cases. Using a dynamic mathematical model of severe acute respiratory syndrome coronavirus 2 (SARS-CoV-2) transmission, we then estimate the impact of reducing case importation on the size of the epidemic in Québec as of July 1^st^, 2020. In addition, we contrast what would have been observed had Québec experienced Ontario’s daily number of imported cases.

## Methods

### Mathematical model

A deterministic SEIR-type of compartmental model of SARS-CoV-2 transmission was developed based on similar frameworks (6,7). The model is semi-mechanistic in the sense that it does not explicitly model interventions. Rather, the impact of those interventions is captured by allowing the reproduction number 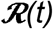 (i.e., average number of successful transmissions per infected person) to vary with time.

In the model (Figure 2), the population is divided into five broad groups: 1) susceptible, 2) exposed (but not yet infectious), 3) infectious and asymptomatic (or pre-clinical), 4) infectious and symptomatic, and 5) removed (i.e., recovered, dead). The duration of the incubation and infectious periods are both assumed to follow an Erlang-2 distribution. The model can be described using ordinary differential equations (Text S1) and the main model parameters are described in Table S1. Whenever appropriate, age-specific parameters are standardized to the age distribution of the Québec population. Cases that have acquired SARS-CoV-2 outside the province are directly imported into the infectious compartments one day before their date of symptoms onset. Since only symptomatic cases could have been detected, we accounted for underreporting of asymptomatic cases by importing infectious individuals in the homonymous compartment, in proportion to the fraction of cases that are asymptomatic. The transmission rate is calibrated and varies with time, reflecting the different interventions implemented (or de-escalated) over time such as case isolation, school closures, and general physical distancing. Specifically, we implemented a time-varying transmission rate, modeled as a first-order random walk with a weekly time step, starting from the implementation of the first physical distancing interventions (March 16^th^, 2020).

**Figure 2.**
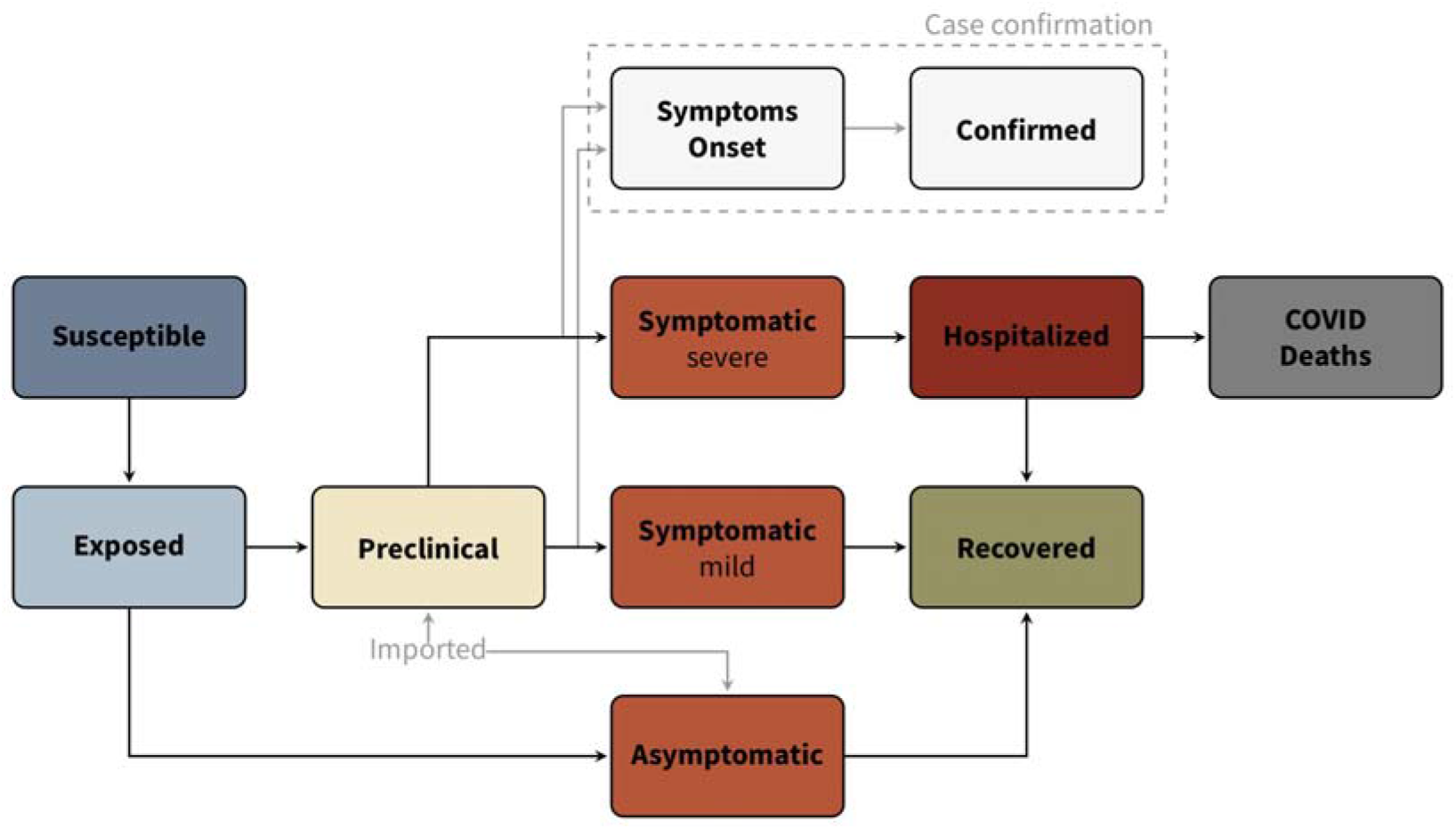
Model structure and inter-compartmental flows of the deterministic SEIR-type model of severe acute respiratory syndrome coronavirus 2 (SARS-CoV-2) transmission.

### Data sources

COVID-19 case surveillance data in Québec is collected for both laboratory confirmed cases and, since March 1^st^ 2020, cases confirmed through epidemiological links. For each confirmed case, basic sociodemographic information, such as age, sex, and region of residence are collected in addition to epidemiological characteristics (date of symptoms onset, likely exposure route, contacts, etc.). For cases with a travel history, date of arrival in Québec and travel destination(s) are also recorded. These data are collected by public health departments and recorded in the “*Fichier V1Ü/COVID-19”*. Hospitalization records are transmitted from each hospital center daily with admission date, transfer and discharge information, and use of intensive care and ventilators. Daily numbers of new COVID-19 hospitalizations are derived from the GESTRED database, before April 6^th^ 2020, and the MED-ÉCHO “Live” database thereafter.

In Ontario, information on COVID-19 cases is reported by each public health unit to the *Ministry of Health and Long-Term Care* through the *Integrated Public Health Information System* (iPHIS). The imported cases in this database lack the date of travel return and, to enable comparisons with Québec, the date of symptoms onset is used.

### Model calibration

Because of important changes over time in testing criteria, confirmation delays in reporting positive cases, COVID-19 testing efforts, and case definition, the model was not fit to the time series of daily confirmed cases. Instead, the model was calibrated to the daily (8) number of new hospitalizations using a negative binomial likelihood to account for overdispersion. We excluded hospital transfers from long-term care facilities (LTCF; named *Centres d’hébergement et de soins longue durée - CHSLD)* as calibration outcomes for two reasons. First, several of these transfers were caused by lack of human resources and/or difficulties to contain outbreaks in these facilities, rather than hospitalizations of individuals requiring acute care. Second, outbreaks in LTCF are epidemiologically distinct from those in the community. The model was initialized on February 27^th^, a day before the first case of COVID-19 was notified in Québec, and where the initial number of infectious individuals was given a uniform prior between 0 and 50. The model’s likelihood and other prior distributions are described in Text S2.

The model is calibrated using Bayesian methods, allowing for efficient propagation of uncertainty. Specifically, we used Markov chain Monte Carlo (MCMC) methods (9) implemented in the *Nimble R* package that allows for fast and flexible Bayesian inferences (10). We used the *Automated Factor Slice Sampler* to sample from the posterior since it performed well at exploring the full parameter space. Convergence was examined using traceplots, and we ensured that the potential scale reduction factor for all parameters and hyperparameters remained close to one (11). The model was coded in R with a C++ back-end (12). The system was solved numerically using a Euler algorithm with a 1.2-hour time step (or 0.05 day). All code is available in a public repository (<u>link here</u>).

### Analyses

After model calibration, we estimated the impact of case importation on SARS-CoV-2’s transmission dynamics by creating counterfactual scenarios where the number of daily imported cases would have been different. This is achieved by sampling the parameter’s posterior distributions, running the model with those samples but altering the number of imported cases, and comparing the counterfactual scenarios of cumulative hospitalizations to the baseline scenario. Specifically, we modeled three counterfactual scenarios. The first corresponds to no imported cases after March 8^th^ (the end of March break in Québec). For this scenario, we kept cases that returned before that date but that had onset of symptoms after March 8th. The second counterfactual scenario compares what would have happened in Québec had the province experienced Ontario’s daily imported case counts. Finally, the third scenario used Québec’s daily imported cases before March 8th but Ontario’s thereafter. For provincial comparisons, imported cases are not adjusted for population sizes since it is the absolute number of importations that affect initial epidemic growth.

### Ethics

Ethical approval for secondary analyses of provincial surveillance data was obtained from McGill University’s *Institutional Review Board*. Secondary analyses of iPHIS data, via the *Ontario COVID-19 Modelling Consensus Table*, were conducted with approval from the University of Toronto Health Sciences’ *Research Ethics Board* (protocol No. 39253).

## Results

From February 28^th^ to March 25^th^, the date at which Québec implemented mandatory quarantine of returning travellers, a total of 1,544 COVID-19 laboratory-confirmed cases had symptom onset among returning travelers to Québec, as compared to 1,150 in Ontario (Figure 3). The majority of those imported cases in Québec were aged 26-65 years and the top-3 countries visited are the United States (24%), France (10%), and Mexico (7%). In Ontario, the top-3 countries visited by travel-related cases are the United States (48%), the United Kingdom (9%), and Mexico (5%).

Almost all imported cases returned to Québec before the mandatory quarantine for travellers with one peak after the main week for March break (March 8th) and another larger peak (March 15^th^) a few days after the *World Health Organization* characterized COVID-19 as a pandemic on March 11^th^ (Figure 3) (13). Compared to Québec, Ontario had slightly lower imported cases (based on dates of symptoms onset) before the mandatory quarantine in that province. The time series of daily number of imported cases for Ontario is flatter than the one in Québec and the peak of the curve was before the end of the March break of Ontario.

**Figure 3.**
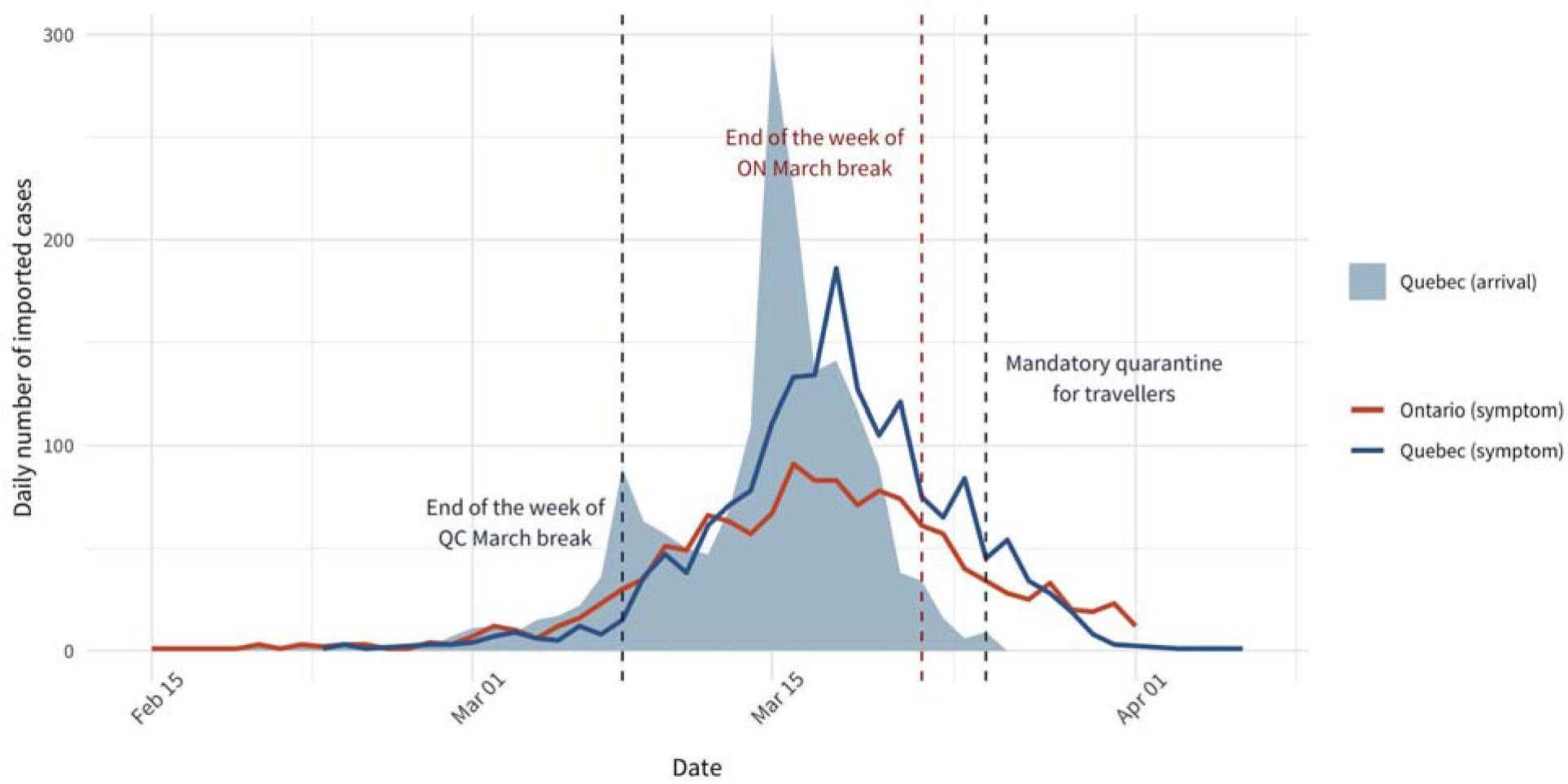
Number of daily imported COVID-19 cases by date of travel return (shaded area; Québec only) and imported cases by date of symptoms onset in Québec and Ontario (lines).

As of July 1^st^ 2020, a total of 6,250 COVID-19 related hospitalizations were recorded in Québec (excluding transfers from LTCF). The daily number of hospitalizations rose rapidly from mid-March to late April, at which point it began to steadily decrease (Figure 4). These dynamics are accurately replicated by the calibrated dynamic model. Measures taken in the early phase of the epidemic, such as cancelling large events, school closure and social distancing, coincide with decreases in the effective reproduction number 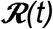 to the unit value within 3-4 weeks of implementation. From mid-April onward, the estimated 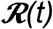 in Québec was below 1.

**Figure 4.**
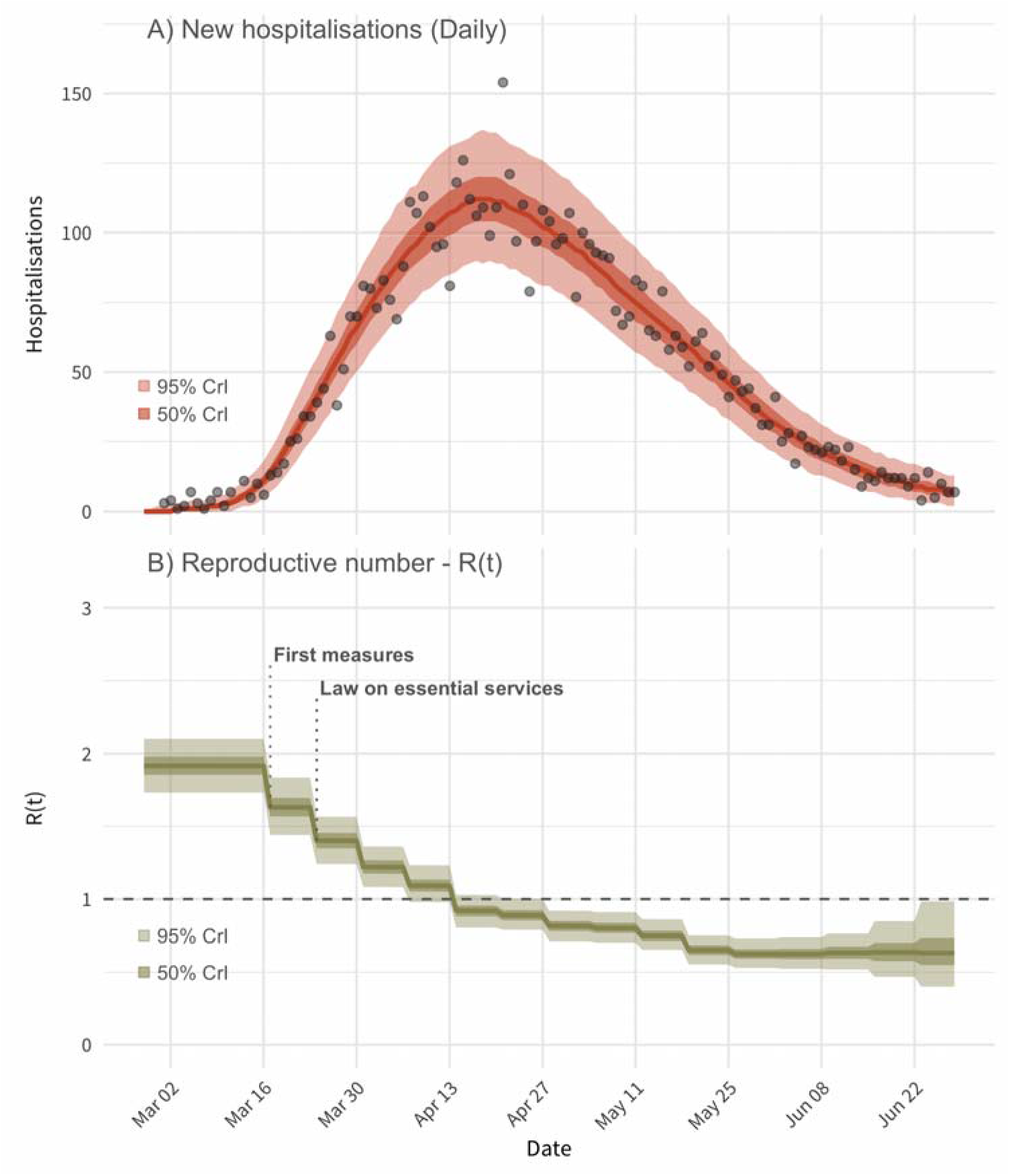
Model fit to the time series of daily hospitalizations in Québec (panel A) with the estimated time-varying effective reproduction number (Panel B). The points in the top panel correspond to the observed number of hospitalizations. In both panels, the solid lines correspond to the median of the model posterior distribution for that outcome and the shaded areas to the 50% and 95% credible intervals.

Results suggest that case importation played an important role in driving the initial spread of SARS-CoV-2 transmission in Québec. Compared to our baseline, using observed daily numbers of imported cases, the counterfactual scenario with no case importation after March 8^th^ resulted in a cumulative number of hospitalizations that would have been 55% lower (95%CrI: 51-59%; Figure 5). However, case importation alone was not sufficient to explain the extent of the epidemic in Québec. Indeed, when using Ontario’s number of imported cases, the cumulative number of hospitalizations was reduced by 12% (95%CrI: 8-16%). If Québec had experienced the same number of daily imported cases as Ontario after March 8^th^, the cumulative number of new hospitalizations would have been 19% lower (95%CrI: 16-23%). Other factors could help explain the Québec-Ontario differential in epidemic size given that Ontario had 29% fewer cumulative hospitalizations (4,423/ 6,250; as of July 5^th^).

**Figure 5.**
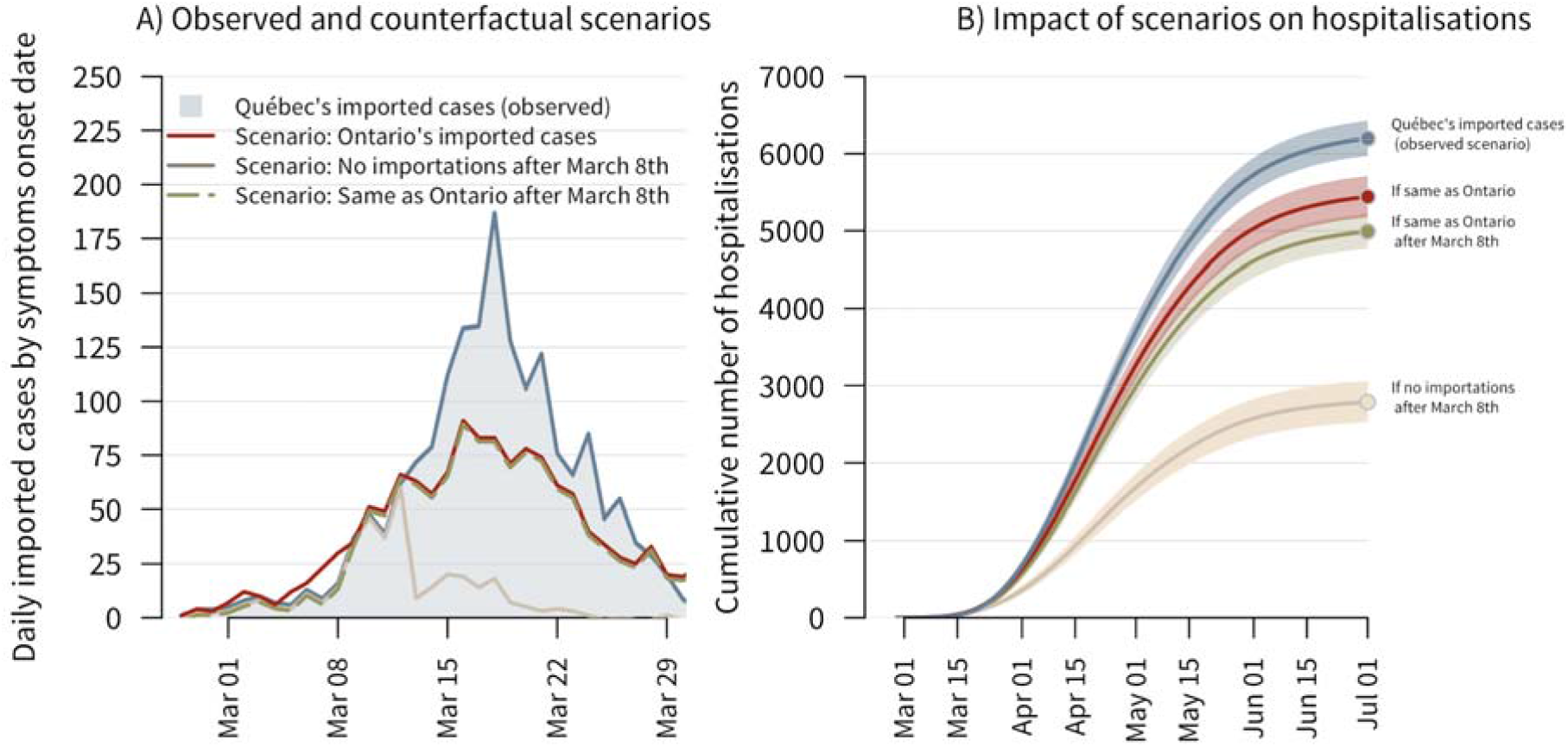
Left Panel (A): observed number of imported cases by date of symptom onsets in Québec (shaded area) and three counterfactual scenarios corresponding to 1) Ontario’s daily number of imported cases, 2) no case importation after March 8th (cases imported before that date can still become infectious later on), and 3) Québec’s observed daily imported cases up to March 8th, after which, Ontario’s daily imported cases are used. For all scenarios, imported cases are assumed to become infectious 1 day prior to symptom onset. Right panel (B): cumulative number of COVID-19 hospitalizations in Québec for the observed scenarios and the three counterfactual scenarios. The solid lines correspond to the median estimates and the shaded areas to the model’s 95% credible intervals.

## Interpretation

This study examined the role of case importation following Québec’s March break in making that province the epicenter of the Canadian epidemic. Using a model of SARS-CoV-2 transmission dynamic, calibrated to detailed surveillance data, we found that cases acquired outside the province played an important role in the initial spread of COVID-19 in Québec. Had Québec not experienced any imported cases after its March break, but otherwise implemented the same interventions, the cumulative number of hospitalizations could have been halved. However, case importation alone is not sufficient to explain why Québec suffered a much more severe epidemic as compared to its close neighbor, Ontario. That province also had many travel-related cases and, had Québec received Ontario’s daily number of imported cases, the epidemic could have been 12-19% smaller in terms of hospitalizations.

The large difference in epidemic sizes of COVID-19 between Québec and Ontario remains puzzling. Health system preparedness and capacity to respond to emerging epidemics could have played a role. For example, Ontario recorded its first SARS-CoV-2 case close to a month before Québec. As such, the largest Canadian province could have better prepared for tracing contacts and isolating imported cases - important non-pharmaceutical interventions to prevent onward transmission (6,7). While public health capacity itself remains difficult to evaluate, Québec’s regional public health agencies *(Directions régionales de la santé publique)* have been subjected to reforms in 2015, with budget decreases of 30% and consequential changes in the organizational structure that could have made coordination more challenging (14,15). Other factors such as suboptimal surveillance systems for early outbreak detections, bottlenecks in testing capacity, initial unavailability of personal protective equipment, and shortages of healthcare personnel, among other things, could have affected transmission in Québec.

These results need to be interpreted considering certain limitations. First, our model mainly reflects transmission in the community and outbreaks occurring in LTCF are not modeled explicitly. As such, it is unclear if the modeled proportional reductions in cumulative hospitalizations for our different scenarios could have directly translated in reduced deaths. Transmission dynamics in LTCF are different, characterized by rapid spread, and healthcare workers moving between LTCF (which happened through the month of May in Québec) could have contributed to onward transmission and high mortality burden in these vulnerable settings, even the number of imported cases would have been lowered. Indeed, although cumulative hospitalizations in Ontario are 29% lower than in Québec, cumulative deaths are 50% lower. Second, the validity of our provincial comparison hinges on the assumption that a comparable fraction of all symptomatic travel-related COVID-19 cases were detected by surveillance systems in both provinces. This assumption could be violated if, for example, more travelers in one province would be returning from countries which were at the time deemed at low risk of COVID-19 (hence less likely to be tested for SARS-CoV-2). Given the reasonably low positivity of 5% before March 25^th^ in Québec, however, incomplete case detection could have been limited. Alternatively, more non-resident tourists from high-risk areas could have visited one province, leading to local chains of transmission, and these tourists would have been missed by case surveillance systems. Assessing levels of SARS-CoV-2 introduction by non-resident tourists is challenging, but we note that Ontario received twice as many non-resident tourists from Europe and the United States during February and March of 2020 (521,081 and 245,521 in Ontario and Québec, respectively), albeit Québec had higher numbers of visitors from France (16,17).

Our analytical framework has several strengths. First, we used detailed surveillance data to inform model development, parameterization, and calibration. Second, the model was calibrated to hospitalization data which is believed to be more robust than case surveillance data, which are affected by time-varying COVID-19 testing efforts, testing protocols, and health-seeking behaviors. Third, we contrasted the experience of Québec and Ontario in terms of imported cases. These two provinces experienced different epidemic trajectories but otherwise share many similarities.

In conclusion, the rapid importation of more than a thousand COVID-19 cases in Québec to date resulted in major outbreaks with more than six thousand cumulative hospitalizations by the beginning of the summer of 2020. Although the size of the Québec epidemic could have been drastically reduced if case importation could have been diminished in early March, comparisons with neighboring Ontario suggests that other factors could explain part of the observed provincial heterogeneity in epidemic size. As Canadian provinces and territories de-escalate their interventions, the potential for SARS-CoV-2 resurgence looms large (18). In this context, it is imperative to implement coordinated strategies that would prevent importation of cases of this magnitude in the future (19). Granular surveillance systems and epidemic intelligence will be key to mitigate SARS-CoV-2 resurgence.

## Data Availability

Part of the data used in this article has not been released in the public domain yet.

https://www.inspq.qc.ca/covid-19/donnees

## Authors’ contributions

DB, AG, MB, MM-G, and YX conceptualized and designed the study; AG, YX, YS, ML, CB, MD, performed data analyses; DD-S, MM-G, ML, AG, and AMS performed model calibration; all authors interpreted the data. AG, MM-G, and YX wrote the first draft of the manuscript and all authors revised it for important intellectual content. All authors have approved the final version for publication and agree to be accountable for all aspects of the work in ensuring that questions related to the accuracy or integrity of any part of the work are appropriately investigated and resolved.

## Acknowledgments

We acknowledge financial support from the *McGill Interdisciplinary Initiative in Infection and Immunity* (MI4; to MM-G), with seed funding from the *MUHC Foundation*, and *a Canadian Institutes of Health Research* (CIHR) *COVID-19 Rapid Research* grant (to SM). MM-G’s research program is supported by a *Canada Research Chair* (Tier 2) in *Population Health Modeling*. SM’s research program is supported by a *Canada Research Chair* (Tier 2) in *Mathematical Modeling and Program Science*. We thank David Landsman (University of Toronto) for helping with data cuts of the Ontario surveillance data.

